# Cohort profile: design, sociodemographic characteristics, chronic disease risk factors, and baseline hypertension and diabetes care cascades of the open, prospective Community-Based chronic disease Care Lesotho (ComBaCaL) cohort

**DOI:** 10.1101/2024.09.18.24313892

**Authors:** Felix Gerber, Giuliana Sanchez-Samaniego, Thesar Tahirsylaj, Thabo Ishmael Lejone, Tristan Lee, Fabian Raeber, Mamakhala Chitja, Malebona Mathulise, Thuso Kabi, Mosoetsi Mokaeane, Malehloa Maphenchane, Manthabiseng Molulela, Makhebe Khomolishoele, Mota Mota, Sesale Masike, Matumaole Bane, Mamoronts’ane Pauline Sematle, Retselisitsoe Makabateng, Ravi Gupta, Irene Ayakaka, Madavida Mphunyane, Lebohang Sao, Mosa Tlahali, Sejojo Phaaroe, Malitaba Litaba, Dave Brian Basler, Kevin Kindler, Pauline Grimm, Eleonora Seelig, Thilo Burkard, Matthias Briel, Frédérique Chammartin, Alain Amstutz, Niklaus Daniel Labhardt

## Abstract

**Purpose:** The open, prospective Community-Based chronic Care Lesotho (ComBaCaL) cohort is the first study to comprehensively investigate socioeconomic indicators, common chronic diseases and their risk factors in a remote rural setting in Lesotho. It serves as a platform for implementing nested trials using the Trials within Cohorts (TwiCs) design to assess community-based chronic care interventions. Here, we present the cohort’s sociodemographic and chronic disease risk factor profile, including self-reported HIV prevalence and hypertension and diabetes care cascades.

**Participants:** Since February 2023, Community Health Worker (CHWs) supported by a clinical decision support and data collection application, have enrolled inhabitants from 103 randomly selected rural villages in Butha-Buthe and Mokhotlong districts in North-East Lesotho. As of May 31, 2024, the cohort includes 5’008 households with 14’735 participants (55% female, median age 19 years). The cohort’s socioeconomic status is low with an International Wealth Index of 26, a monthly household income of 42.4 USD and low levels of formal education. Among the 7’917 adult participants, 42.5% are overweight or obese, with higher rates among women, and 33.1% smoke tobacco, with higher rates among men. Self-reported HIV prevalence is 15.1% with a 98.4% treatment rate. Hypertension prevalence is 17% with a 56% control rate and diabetes prevalence is 4% with a 39% control rate.

**Findings to date:** The cohort’s low socioeconomic status is linked to multiple health risks including insufficient access to clean energy, essential healthcare services, adequate sanitary facilities and secure food supply. Besides the expected high HIV prevalence, we found significant hypertension, diabetes and cardiovascular risk factor prevalences. While treatment and control rates for diabetes and hypertension are higher than in similar settings, they remain below global targets.

**Future plans:** Ongoing cluster-randomized TwiCs, which will be completed in 2025, are assessing the effectiveness of community-based, CHW-led care interventions for diabetes and hypertension. CHWs will continue to closely monitor the cohort and integrate additional measurements such as HIV testing. This will provide further insights into the dynamics and interactions of chronic diseases and inform the development of future nested trials on innovative community-based prevention and care interventions.

**Registration:** NCT05596773

**Strengths and limitations:**

– Comprehensive Data Collection: The ComBaCaL cohort offers comprehensive data on sociodemographics, chronic disease risk factors, and hypertension and diabetes care cascades within a large, representative sample of the rural population in Lesotho.
– Community-Based Approach: Data is captured by local Community Health Workers residing in the study villages using a tailored clinical decision support and data collection application. This approach allows for continuous data collection, remote monitoring by study staff, and data verification, ensuring nearly complete village enrolment and high data quality.
– Efficient study design: The cohort utilizes the Trials within Cohorts (TwiCs) design, which allows for the efficient implementation of multiple randomized nested trials to assess the effectiveness of innovative health interventions.
– Reliance on self-reported data: Assessments other than hypertension and diabetes screening outcomes rely on self-reported data, which may have limited correlation with objective assessments.
– Limitations in data scope: Clinical data on chronic conditions other than hypertension and diabetes remain limited and anthropometric and behavioural risk factor data for children has not yet been collected.

## Introduction

Globally, chronic non-communicable diseases (NCDs) are the leading cause of death and disability with a particularly high burden in low- and middle-income countries [1]. In Africa, the NCD burden has risen significantly over the past two decades, driven by the increasing prevalence of lifestyle risk factors such as unhealthy diet, smoking, insufficient physical activity, obesity, and exposure to air pollution [2,3]. Concurrently, the burden associated with human immunodeficiency virus (HIV), a main cause of morbidity and mortality in the region, is decreasing due to the widespread roll-out of effective antiretroviral therapy. As a result, projections indicate that by 2030, NCDs will surpass communicable, maternal, neonatal, and nutritional diseases combined as the leading cause of mortality in Southern Africa [4]. This epidemiological transition signifies a critical need for enhanced chronic care services in the region, driven by both the rising NCD prevalence and the transformation of HIV into a manageable chronic illness [5,6]. To meet this challenge, Southern African health systems must undergo substantial transformation, necessitating concerted research efforts to facilitate this process [7]. Moreover, there are intricate social and biomedical interactions between HIV and NCDs and both are mediated by shared risk factors which are closely linked to socioeconomic development highlighting the complexities of effective responses [7–9]. However, epidemiological data on chronic diseases, particularly in rural low-resources settings, remains scarce, with only very few cohort studies or trials examining HIV, NCDs, and socioeconomic indicators together [10]. Additionally, further health systems research is essential to explore how integrated chronic care can be developed within resilient health systems capable of concurrently addressing NCDs and HIV [7,9,11].

A promising approach to increase the chronic care capacities of health systems with limited professional workforce and financial resources is the decentralization of healthcare services with task-shifting to lay Community Health Workers (CHWs) [12–16]. The strategic integration of community-based healthcare services in existing health systems is supported by the World Health Organization (WHO), and the United Nations Programme on HIV/AIDS (UNAIDS) is promoting scale-up of CHW-delivered services in Africa [17,18]. CHWs bring services closer to the community, reduce access barriers such as transport costs, travel time and low awareness, and may offer more equitable and less stigmatized access to health services than facility-based care. In addition, CHW systems have the potential to strengthen civil society and create job opportunities in rural areas [17].

Although various modelling studies suggest that integrated community-based chronic care may be cost-effective in Southern Africa, robust evidence around community HIV/NCD delivery platforms and their key enablers is missing [19–23]. Evidence from implementation research in resource-limited settings on primary healthcare in rural areas including pharmacological interventions is scarce [24]. For southern Africa, no evidence on the clinical effectiveness of community-based NCD or integrated chronic care models is available [21,22].

Lesotho is an example of an African lower-middle-income country where the burden of NCDs is rapidly increasing while HIV prevalence remains high and the population of people living with HIV is ageing [25,26]. The Community-Based chronic Care Lesotho (ComBaCaL) cohort study provides detailed socioeconomic information and epidemiological data on the two most common chronic NCDs, arterial hypertension and diabetes, and on behavioral risk factors in rural Lesotho, a remote setting with high HIV prevalence. Furthermore, it will serve as a platform for the assessment of novel community-based chronic care models in nested trials using the Trials within cohorts (TwiCs) design, that will generate evidence about how CHWs could support the development of a resilient health system able to provide widespread equitable access to chronic care.

Here we present the baseline characteristics of the ComBaCaL cohort population including socioeconomic characteristics, lifestyle risk factors, and diabetes, hypertension and self-reported HIV prevalence, and treatment status. These findings contribute significantly to our understanding of the health needs in rural Lesotho and inform the development of effective, community-based chronic care interventions.

## Cohort description

### Setting

The ComBaCaL cohort study is situated in Butha-Buthe and Mokhotlong districts in north-east Lesotho, a landlocked, mountainous country in southern Africa with an estimated population of about 2.3 million inhabitants [27]. The majority of the population lives in rural areas with difficult access to healthcare facilities [28]. Both districts feature one central town with surrounding rural areas with remote villages and poor transport infrastructure. In the two districts combined, three physician-led secondary hospitals and 19 nurse-led primary-level health centres serve a population of approximately 220’000 people [29].

At 19%, Lesotho has the second-highest adult HIV prevalence globally [30]. A recent population-based survey in Butha-Buthe and Mokhotlong found adult prevalences of 21.6% and 5.3% for arterial hypertension and diabetes [25] with suboptimal treatment and control rates and high rates of end-organ disease [31,32].

As in many other African countries, the Lesotho health system faces significant funding challenges [33]. The country faces a shortage of medical professionals, with just 20.7 doctors and nurses per 10’000 people. This is less than 50% of the WHO defined threshold deemed necessary to make progress towards universal health coverage [34]. Nevertheless, Lesotho has managed to reduce HIV transmission and mortality considerably over the last years. This success is largely attributed to a decentralized HIV testing and care system that effectively leverages lower cadre healthcare workers and CHWs, called Village Health Workers in Lesotho, to deliver accessible and equitable services for the urban and rural population alike. The integration of lay CHWs into the health system structures has been adopted in Lesotho since 1978 [35,36]. CHWs are considered lay volunteers in Lesotho but they receive a monthly government stipend of roughly 45 USD [37]. Currently, CHWs the community-based health care delivery is focused on HIV and maternal and neonatal diseases. NCD care is only provided at the health facilities while the high prevalence of NCDs and relevant awareness, diagnosis and treatment gaps underscore the need to develop more accessible NCD service models [25,31,32]. The Lesotho CHW program represents a meaningful starting point for implementation research on community-based chronic care models as it is representative for the health systems in many other African countries, where lay worker-led care has a similar standing.

### Objectives and Design

The ComBaCaL cohort is a prospective research and service delivery platform, with the aim to assess the prevalence, burden, and risk factors of chronic diseases, especially arterial hypertension, type 2 diabetes, and HIV in rural Lesotho. In addition, the ComBaCaL cohort serves as a platform for the assessment of community-based, CHW-led chronic care interventions in nested randomized trials using a TwiCs approach [38–40]. TwiCs is a pragmatic randomized design [41] with the potential to overcome some challenges of traditional randomized trials like slow recruitment, limited external validity, burdensome consent procedures, or undesirable study-related behaviour of participants (‘disappointment effects’) [39,40,42]. At enrolment, participants not only consent for regular cohort data collection, but also for potential randomization into future nested trials assessing low-risk interventions. If randomized to the intervention arm of a trial, participants are offered the intervention and have then the option to accept or decline it. If randomized to the control arm of the trial, participants are not contacted but continue usual care in the cohort [43]. In the ComBaCaL cohort, three cluster-randomized TwiCs assessing CHW-led care for hypertension and diabetes are ongoing at the time of submission and expected to be completed in 2025 [44,45].

### Sampling of villages and recruitment of CHWs

Based on the 2016 Lesotho Population and Housing Census [29], a total of 675 village-clusters were identified in the rural areas of Mokhotlong (321) and Butha-Buthe (354) districts. A few villages located in neighbouring districts close to district borders and within the catchment areas of Mokhotlong or Butha-Buthe health facilities were also included and assigned to Mokhotlong or Butha-Buthe district depending on the location of the associated health facility. Out of the 675 villages, 70 (35 per district) were excluded because they were involved in other chronic disease related studies. Out of the 605 remaining villages, 140 (70 per district) were randomly sampled, stratified by district and access to health facility (easy versus difficult access, defined as needing to cross a mountain or river or travel >10 km to the nearest health facility), by a statistician not involved in the study. The 140 villages were assessed for eligibility applying the following criteria: estimated village size of 40 to 100 households, geographic distribution of households that allows coverage by one CHW, village consent obtained from village chief, and possibility to identify or recruit a CHW from the village population. The upper limit of household numbers was chosen to ensure each study village could be served by one CHW. The lower limit of household numbers was chosen to ensure a reasonable minimal workload per CHW and to minimize the number of empty clusters for nested trials enrolling disease-specific subpopulations. The village eligibility assessment was conducted after the random sampling because of its operational complexities requiring physical visits of the study team and consent by the village chiefs, which was not feasible to conduct for the entire sampling pool of 605 villages. During the assessment, 29 villages were excluded (23 had less than 40 households, 4 had more than 100 households, 2 consisted of smaller sub-clusters that were too far apart to be served by one CHW). See figure 1 for the village selection flow.

**Figure 1.**
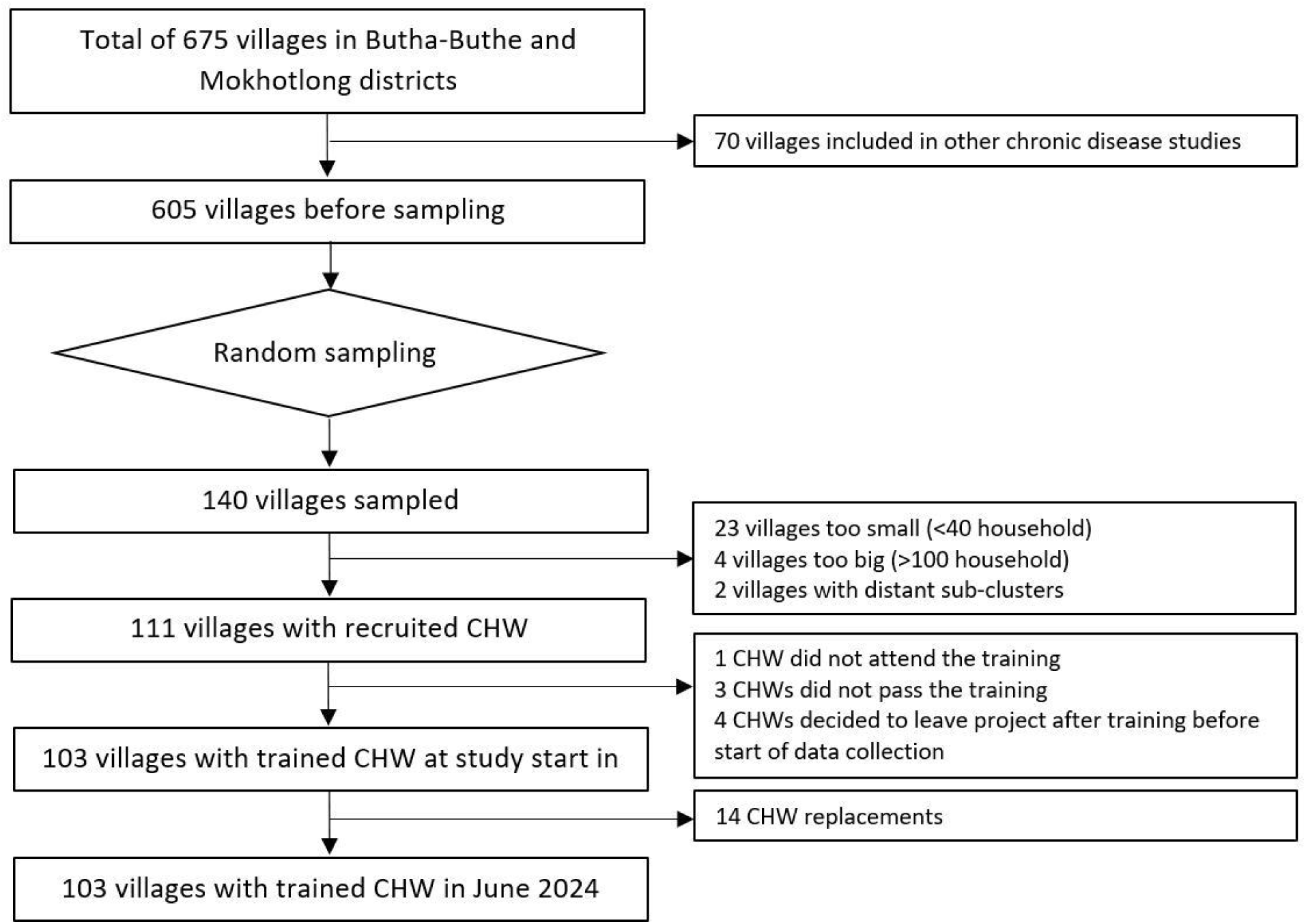
Selection of villages and CHWs. CHW: Community Health Worker

Criteria for CHW selection were in line with the Lesotho Village Health Program Policy [37] and included the following: primary residence in the village, record of trustworthiness and ability to maintain confidentiality (according to village chief), aged between 20 and 50 years, being able to provide written reports and to do basic mathematical calculations, having at least an educational level equivalent to a high school leaving certificate (Junior Certificate), having good social and communication skills, and being able to speak, understand and write in English. In villages where an existing CHW fulfilled the criteria, he/she was invited to participate in the project. In villages where there was no CHW or where the existing CHW did not fulfil the criteria, a new CHW was elected by the village population during a community gathering after a pre-selection of candidates by the village chief, the study team and the district health management team of the Ministry of Health.

CHWs from 111 villages meeting the eligibility criteria were invited to an initial 10-day training on consent procedures, recognition and reporting of clinical events, and data collection. 103 CHWs passed the training and started the participant enrolment in their villages in February 2023 (1 did not attend the training, 3 did not pass the training, and 4 decided not to participate after the training). Villages are distributed in the rural catchment areas of all 22 health facilities in the two districts (see figure 2). One Mokhotlong village is located in Thaba-Tseka district close to the district border but falls under the catchment area of a Mokhotlong health centre. Between February 2023 and June 2024, CHWs in 14 villages had to be replaced (5 got a permanent job outside the village, 1 went to university, 3 got elected as local councillors, 5 were unable to perform the tasks due to lack of time, motivation or skills). If CHWs needed to be replaced, a new CHW was elected by the community in collaboration with district health authorities and trained to ensure that service delivery and data collection in the villages remained uninterrupted.

**Figure 2.**
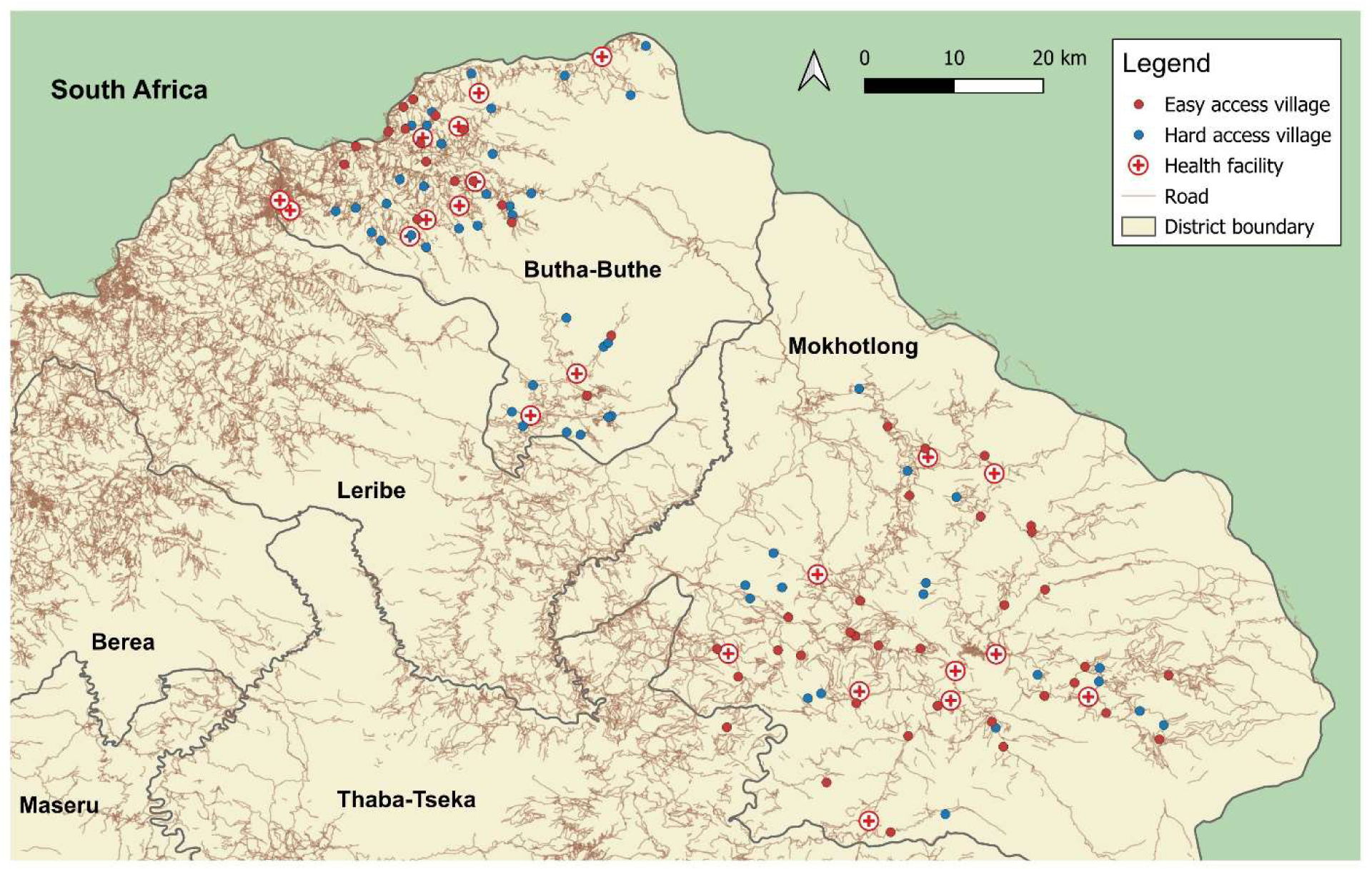
Location of the study villages and health facilities in Butha-Buthe and Mokhotlong districts in Lesotho. Hard access village: Needing to cross a mountain or river or travel >10 km to the nearest health facility. Map was produced by TL using QGIS v3.34 [46].

### Eligibility, consent and enrolment

Oral consent from the village chiefs was obtained during village eligibility assessment prior to the CHW training with all approached village chiefs giving consent for their village to participate. All inhabitants of the selected villages independent of age or any other factors are eligible for participation in the ComBaCaL cohort.

Each cohort village is managed by one CHW, who lives in the village and is supported by a tablet-based application for consent documentation, data collection, and clinical decision support (ComBaCaL app) [47,48]. For participant enrolment, CHWs visit all households in their villages. Prior to individual enrolment, households are registered, and oral household consent asked from the household head or a representative.

After obtaining household consent, present household members are asked for individual written consent using a tablet-based consent application. The number of absent household members is registered and the CHW returns to households with absent members in the next days to complete recruitment. Based on participants’ characteristics (age, literacy, need for a guardian), the consent application displays appropriate simplified study information and allows for electronic signature on the informed consent form. Illiterate participants confirm informed consent by drawing a cross in the electronic signature field, countersigned by an impartial witness. For adolescents aged 10 to 17 years, written consent is sought from a guardian together with written assent from the adolescent. For children below 10 years, written consent is sought from a guardian without assent from the child. Signed consent forms are uploaded in the study database of the ComBaCaL app and checked for correctness and completeness by the study team remotely and reconsent tasks are triggered via the ComBaCaL app in case of errors.

Following the French consent pattern of the TwiCs approach [40], participants consent to being randomized as part of future TwiCs together with consent for ComBaCaL cohort data collection. The consent form is available on clinicaltrials.gov (NCT05596773). Consent can be withdrawn any time on individual or household level. Pseudo-anonymized data collected until the time of withdrawal is retained in the database.

The ComBaCaL cohort is an open cohort and new inhabitants of the selected villages are approached for consent as soon as the CHW becomes aware of the new person moving into the village or being born. After death or emigration, the participant is removed from the active cohort population while the collected data remains in the database. Participants who emigrated, and then move back into the village are reactivated and continue regular follow-up. The presence of the CHWs in the villages and the continuous enrolment of new participants, deactivation of leaving participants, and reactivation of returning participants ensures that the cohort population is constantly aligned with the actual number of consenting people staying in the study villages.

### Measurements

All data are collected by CHWs during household visits using the tailored ComBaCaL application that provides algorithmic clinical decision support and serves as data collection tool. In addition to the initial 10-day training on consent procedures, recognition of clinical alarm signs and symptoms and relevant skills for data collection, CHWs were trained on diabetes and hypertension screening and diagnosis separately during two additional 3-day trainings.

For consenting households, household level data including socioeconomic indicators and GPS coordinates are collected. Household wealth is assessed using the Demographic and Health Survey (DHS) Program wealth index questions for Lesotho [49]. The DHS wealth index is a 15-item questionnaire that assesses household assets and utility services, including country-specific assets that are viewed as indicators of economic status [50]. The DHS questionnaire is used to calculate the International Wealth Index (IWI). The IWI, an asset-based index, facilitates comparisons of household economic status across low- and middle-income countries. It operates on a scale from 0 to 100, where 0 represents the lowest housing quality and absence of essential consumer durables, while 100 signifies the highest housing quality and ownership of all relevant consumer durables [51]. Previous research has demonstrated robust correlation between the IWI and other health and socioeconomic indicators [51,52].

Food security is assessed using questions from the Household Food Insecurity Access Scale (HFIAS) [53]. Only six out of the nine questions included in the full HFIAS are asked. Therefore, the HFIAS score is not calculated, but food insecurity categories based on the highest-ranked answer to the available six questions are presented [53]. Furthermore, access to healthcare is assessed using validated questions about whether household members could not access required healthcare services and medications in the last 12 months [54]. For all participants, age, sex, current or previous tuberculosis diagnosis, and self-reported HIV status and intake of antiretroviral therapy for those reporting to live with HIV are collected. For adolescents (10-17 years) and adults (≥18 years), tobacco and alcohol consumption, salt, fruit and vegetable consumption using the WHO STEPS instrument [55], consumption of unhealthy food items, beverages and meat using a food frequency questionnaire adapted from an assessment tool for obesity used in South Africa [56], physical activity using the validated International Physical Activity Questionnaire Short Form (IPAQ-SF) [57], weight, height, and abdominal circumference are collected. For the assessment of unhealthy food items and beverages consumption, the numbers of days per week on which the included items were consumed are asked. The food items included are the most frequently consumed unhealthy food items based on the judgement of the local study team members and are not validated. For analysis, the items are categorized in sweet items (candy, chocolate and biscuits), fried high-carbohydrate items (crisps, fries, fat cakes (traditional fried bread buns)) and sweet beverages (fruit juices, carbonated soft drinks). For each category the highest frequency of consumption of any of the included items is reported. For analysis of physical activity, we calculate the metabolic equivalent of task (MET) minutes per week and categorize in low, moderate and high physical activity according to the IPAQ scoring protocol [57]. Only age, sex and overall number of children and adolescents are included in this manuscript as part of the ComBaCaL cohort overview, while further data on children and adolescents will be published separately.

For adults aged 18 years and above, awareness and history of hypertension and diabetes, diagnosis of relevant chronic diseases and complications (myocardial infarction, stroke, heart failure, chronic kidney disease, vision impairment, peripheral arterial disease, peripheral neuropathy, asthma, chronic obstructive pulmonary disease (COPD), diabetic foot syndrome), and current medication intake are documented. For the documentation of clinical information, self-reports and the participants’ personal health booklets are used. CHWs are guided during the data collection by questions and additional explanatory notes displayed in the ComBaCaL app in English and Sesotho, the local language.

Adult participants aged 18 years or older are screened for hypertension according to the diagnostic algorithm of the Lesotho Standard Treatment Guidelines [58]. Blood pressure measurements are conducted using automated machines (Omron M3 Comfort [HEM7131-E] [59]). Measurements are taken after determination of the correct cuff size in a sitting position after five minutes of rest with feet on the floor, the arm supported without talking or moving during the measurement. At the first visit, the reference arm is determined by measuring blood pressure on both arms. The arm with higher systolic blood pressure is identified as reference arm and used for all subsequent measurements. The blood pressure value is calculated as the mean of the last two out of three consecutive measurements at intervals of one minute. For the diagnosis of hypertension, two elevated measurements in the range of 140-179/90-109 mmHg on two different days are required or two measurements of 180/110 mmHg or higher on the same day, at least 30 minutes apart. Participants reporting intake of antihypertensive medication or newly diagnosed according to the diagnostic criteria are considered as having hypertension. Hypertension diagnosis awareness is defined as reporting having been previously diagnosed with hypertension. Hypertension treatment control is defined as having a blood pressure below 140/90 mmHg.

Adult participants aged 40 years or older or having a BMI of 25 kg/m^2^ or higher or reporting intake of antidiabetic medication are screened for diabetes according to the diagnostic algorithm of the Lesotho Standard Treatment Guidelines [58] using handheld glucometers for capillary blood sugar measurements. Diagnostic criteria include a random blood sugar of ≥11.1 mmol/l or a fasting blood sugar ≥7.0 mmol/l either in presence of cardinal diabetes symptoms (polyuria, polydipsia, and weight loss) or after previously elevated blood sugar (fasting or random blood sugar ≥5.6 mmol/l) on a different day. Participants reporting intake of antidiabetic medication or newly diagnosed according to the diagnostic criteria are considered as having diabetes. For people diagnosed with diabetes based on blood glucose values, glycated haemoglobin (HbA1c) is measured. Diabetes diagnosis awareness is defined as having been previously diagnosed with diabetes. Diabetes treatment control is defined as having a fasting blood glucose below 7.0 mmol/l or a HbA1c below 6.5% if fasting blood glucose is missing.

### Data management and analysis

All CHWs received a password-protected tablet with the ComBaCaL app installed. The ComBaCaL app is based on the Community Health Toolkit Core Framework, a widely used, offline-first, open-source software toolkit designed for community health systems [60].

Data are collected in real time on the tablets and synchronized regularly to a secure server hosted at the University Hospital Basel, Switzerland. All shared data exports are pseudonymized. Data are continuously monitored locally by the CHW supervisors and centrally by the data management team of the University Hospital Basel with CHW supervisors contacting the CHWs through phone calls or field visits in case of potential data errors for verification.

Descriptive statistics are used to present participant characteristics. Missing data are reported as missing in all tables. All analyses are done using R core team, version 4.3.1. 2023 [61].

## Participants’ characteristics

Since February 2023, in the 103 study villages, 5’274 households with 16’461 people were approached for household consent to participate in the ComBaCaL cohort. Through repeated home visits by the CHWs, nearly all households of the 103 villages were reached. For 11 households (0.2%) with 31 people, household consent was refused. 16’430 individuals from 5’263 households were approached for individual consent. 16’158 participants (98.3% individual consent rate) consented to participation. Of those, up to June 2024, 189 withdrew consent (1.1%), 113 died (0.7%), 1’119 moved out of the village (8.7%), and 2 did not yet have baseline data collected (0.03%). Overall, in June 2024, the ComBaCaL cohort comprised 14’735 active participants with available baseline information from 5’008 households (see figure 3).

**Figure 3.**
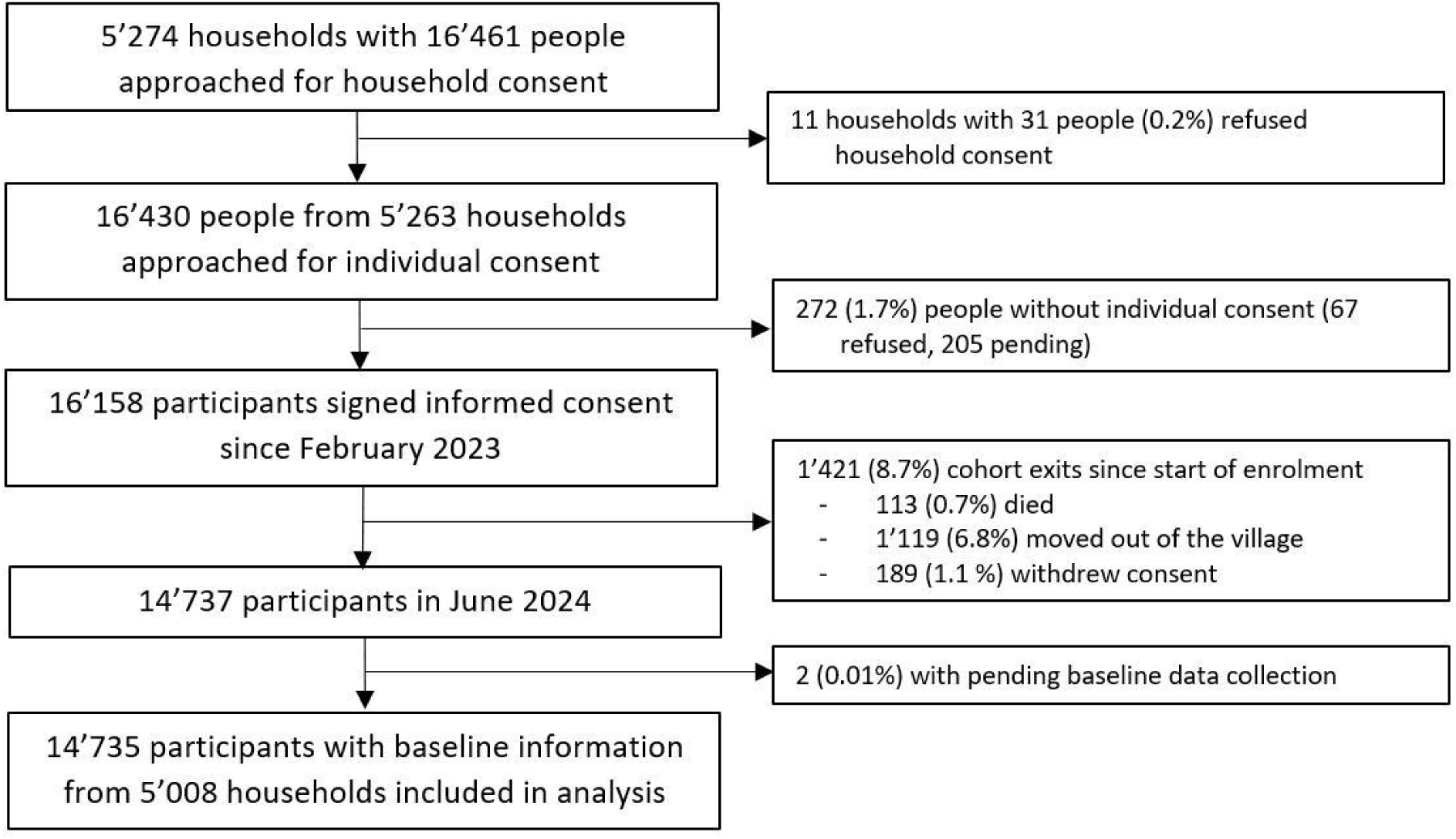
Overview of consent, enrolment, and retention of households and participants in the ComBaCaL cohort.

Number of participants per village range from 58 to 302 with a median of 140.

Table 1 provides an overview of socioeconomic indicators at household level, the complete household-level data is available in the supplement. The median asset-based IWI score and monetary monthly household income are 26 (IQR 17, 36) and 42.4 USD (IQR 18.6, 63.6 USD), respectively. In a typical ComBaCaL cohort household, there are three people living (IQR 2, 4) in one or two bedrooms. Most houses are made of mud walls (61.9%) with a natural straw roof (61.9%). The main source of drinking water is a community tap (81.4%), and toilet facilities mostly consist of pit latrines (47.6%) while 39.1% of households do not have a toilet and defecate in fields or bushes. Only 13.2% of households have access to electricity. The most widely used cooking and heating fuels are wood (84.1% for cooking and 63.9% for heating) and animal dung (6.0% for cooking and 26.4% for heating). Roughly half of households (50.9%) have geographically difficult access to a health facility as per the definition of living more than ten kilometres away from the closest health facility or having to cross a river or a mountain to reach the facility. 22.4% of households reported that at least one member was unable to access healthcare services in the past 12 months despite needing them, while 16.4% faced difficulties obtaining required medications. The main reasons stated for the difficulties in accessing healthcare services were lack of money or difficulties to get transportation. 62.4% of households own agricultural land and for 24.2% of households, farming is the main source of income.

**Table 1.** Household level socioeconomic indicators. IQR: Interquartile range.

| Characteristic | N = 5,008<sup>1</sup> |
| --- | --- |
| **Number of household members, median (IQR)** | 3 (2, 4) |
| **International Wealth Index, median (IQR)** | 26 (17, 36) |
| **Monthly household income (USD)<sup>1</sup>, median (IQR)** | 42.4 (18.6, 63.6) |
| Unknown / refused to say | 7 |
| **Main source of household income** |  |
| Salary / wages<sup>2</sup> | 1,289 (25.7%) |
| Informal business (e.g. street vendor) | 1,109 (22.1%) |
| Formal business (e.g. retail trade) | 37 (0.7%) |
| Farming | 1,213 (24.2%) |
| Rentals/interest | 61 (1.2%) |
| Grants/pension | 1,109 (22.1%) |
| Unknown / refused to say | 190 (3.8%) |
| **Household Food Insecurity Access category** |  |
| Food secure | 2,385 (47.6%) |
| Mildly food insecure | 167 (3.3%) |
| Moderately food insecure | 965 (19.3%) |
| Severely food insecure | 1,489 (29.7%) |
| Unknown / refused to say | 2 |
| **Geographically hard access to health facility<sup>3</sup>** | 2,548 (50.9%) |
| **Difficulties accessing health care<sup>4</sup>** | 1,120 (22.4%) |
| **Difficulties accessing medications<sup>5</sup>** | 821 (16.4%) |
| **Drinking water** |  |
| Running (piped) water inside dwelling or yard | 79 (1.6%) |
| Neighbour's tap | 43 (0.9%) |
| Public / community tap | 4,075 (81.4%) |
| Borehole | 548 (10.9%) |
| Dam / river / rainwater | 257 (5.1%) |
| Unknown / refused to say | 6 (0.1%) |
| **Toilet** |  |
| Flush toilet | 33 (0.6%) |
| Ventilated improved pit latrine | 628 (12.5%) |
| Non-improved pit latrine | 2,381 (47.6%) |
| None / bush / field | 1,959 (39.1%) |
| Unknown / refused to say | 7 (0.1%) |
| **Electricity available** | 663 (13.2%) |
| **Cooking fuel** |  |
| Wood | 4,210 (84.1%) |
| Animal dung / straw / grass | 302 (6.0%) |
| Gas | 237 (4.8%) |
| Paraffin | 147 (2.9%) |
| Electricity | 106 (2.1%) |
| Other / unknown / refused to say | 6 (0.1%) |
| **Heating fuel** |  |
| Wood | 3,198 (63.9%) |
| Animal dung / crop waste | 1,321 (26.4%) |
| Gas | 41 (0.8%) |
| Paraffin | 322 (6.4%) |
| Electricity | 68 (1.4%) |
| Other / unknown / refused to say | 57 (1.2%) |
| **People per bedroom** |  |
| 1-2 | 2,527 (50.5%) |
| 3-4 | 1,880 (37.5%) |
| 5 or more | 599 (12%) |
| Unknown / refused to say | 2 (<0.1%) |
| **Wall material** |  |
| Mud | 3,099 (61.9%) |
| Mud and cement | 1,042 (20.8%) |
| Corrugated iron | 230 (4.6%) |
| Bare brick | 245 (4.9%) |
| Finished/plaster | 348 (6.9%) |
| Other / unknown / refused to say | 44 (0.8%) |
| **Roof material** |  |
| Natural | 3,099 (61.9%) |
| Rudimentary (wood planks, cardboard) | 242 (4.8%) |
| Finished (metal, wood, tiles) | 1,548 (30.9%) |
| Other / unknown / refused to say | 119 (2.4%) |
| **Agricultural land ownership** | 3,127 (62.4%) |
| <sup>1</sup> Collected in Lesotho Loti, converted using the bilateral conversion rate on May 31, 2024 [62] |  |
| <sup>2</sup> Salaries and wages of household members or other people (i.e. of family members living outside the household) |  |
| <sup>3</sup> Defined as needing to cross a mountain or river or travel >10 km to the nearest health facility. |  |
| <sup>4</sup> Defined as at least one household member not being able to consult a healthcare provider despite needing it during the last 12 months. |  |
| <sup>5</sup> Defined as at least one household member experiencing difficulties obtaining required medication. |  |

Table 2 provides an overview of the individual-level socioeconomic characteristics of the adult participants enrolled in the ComBaCaL cohort by May 31, 2024, the complete data is available in the supplement. Median age of participants is 19 years (interquartile range (IQR) 9 and 42 years) with a higher median age of female participants (21 versus 17 years). 47% of participants are below 18 years. 69.4% do not have any formal or only primary level education with more males having no education (20.7% versus 6.1%). 12.9% of adult participants work for pay (17.3% of male and 9.7% of female) while 37.5% are self-employed or working in agriculture (53.8% of male and 25.1% of female). 38% of female and 5.6% of male participants are homemakers.

**Table 2.** Socioeconomic characteristics of ComBaCaL cohort participants. IQR: Interquartile range.

| Characteristic | Female, n (%),<br>8064 (55%) | Male, n (%), 6671<br>(45%) | Total, n (%),<br>14735 (100%) |
| --- | --- | --- | --- |
| **Age (years), median (IQR)** | 21 (10, 45) | 17 (8, 39) | 19 (9, 42) |
| <10 years | 1,936 (24.0%) | 2,006 (30.1%) | 3,942 (26.8%) |
| 10-17 years | 1,568 (19.4%) | 1,351 (20.3%) | 2,919 (19.8%) |
| 18-39 years | 2,148 (26.6%) | 1,662 (24.9%) | 3,810 (25.9%) |
| 40-64 years | 1,529 (19.0%) | 1,185 (17.8%) | 2,714 (18.4%) |
| ≥65 years | 883 (10.9%) | 467 (7.0%) | 1,350 (9.2%) |
| **District** |  |  |  |
| Butha-Buthe | 4,002 (49.6%) | 3,393 (50.9%) | 7,395 (50.2%) |
| Mokhotlong | 4,062 (50.4%) | 3,278 (49.1%) | 7,340 (49.8%) |
| **Adult participants ≥ 18 years** | 4,560 (56.5%) | 3,314 (49.7%) | 7,874 (53.4%) |
| **Relationship status** |  |  |  |
| Single/divorced/widowed | 1,861 (40.8%) | 1,270 (38.3%) | 3,131 (39.8%) |
| Married/committed relationship | 2,690 (59.0%) | 2,024 (61.1%) | 4,714 (59.9%) |
| Unknown/refused to say | 9 (0.2%) | 20 (0.6%) | 29 (0.4%) |
| **Education** |  |  |  |
| No schooling | 276 (6.1%) | 684 (20.7%) | 960 (12.2%) |
| Primary school | 2,623 (57.5%) | 1,879 (56.7%) | 4,502 (57.2%) |
| Secondary/high school | 1,448 (31.8%) | 624 (18.8%) | 2,072 (26.3%) |
| Tertiary/higher/post high school | 209 (4.6%) | 121 (3.7%) | 330 (4.2%) |
| Unknown/refused to say | 3 (0.1%) | 4 (0.1%) | 7 (0.1%) |
| **Work status** |  |  |  |
| Working for pay | 444 (9.7%) | 572 (17.3%) | 1,016 (12.9%) |
| Self-employed | 703 (15.4%) | 709 (21.4%) | 1,412 (17.9%) |
| Working on own plot or looking after livestock | 469 (10.3%) | 1,075 (32.4%) | 1,544 (19.6%) |
| Helping another family member with their business, without pay | 33 (0.7%) | 30 (0.9%) | 63 (0.8%) |
| Full-time student | 90 (2.0%) | 38 (1.1%) | 128 (1.6%) |
| Homemaker (looking after children/others/home) | 1,734 (38.0%) | 184 (5.6%) | 1,918 (24.4%) |
| Long term sick or disabled | 53 (1.2%) | 41 (1.2%) | 94 (1.2%) |
| Retired | 205 (4.5%) | 136 (4.1%) | 341 (4.3%) |
| Unemployed | 702 (15.4%) | 461 (13.9%) | 1,163 (14.8%) |
| Unknown/refused to say | 127 (2.8%) | 68 (2.1%) | 195 (2.5%) |

### HIV status, chronic disease complications, and risk factors

Table 3 shows the overview of self-reported HIV and ART status alongside chronic disease risk factors and complications. Further details are in supplement.

**Table 3.** Body mass index, behavioural cardiovascular risk factors, self-reported HIV and ART status of participants. BMI: Body-mass-index, COPD: Chronic Obstructive Pulmonary disease.

| Characteristic | Female, n(%),<br>4560 (58%) | Male, n(%),<br>3314 (42%) | Total, n(%),<br>7874 (100%) |
| --- | --- | --- | --- |
| **Self-reported HIV status** |  |  |  |
| HIV positive | 782 (17.1%) | 406 (12.3%) | 1,188 (15.1%) |
| Tested negative ≤ 12 months ago | 2,435 (53.4%) | 1,503 (45.4%) | 3,938 (50.0%) |
| Tested negative > 12 months ago | 846 (18.6%) | 684 (20.6%) | 1,530 (19.4%) |
| Unknown | 453 (9.9%) | 695 (21.0%) | 1,148 (14.6%) |
| Refused to say | 44 (1.0%) | 26 (0.8%) | 70 (0.9%) |
| **ART intake among those HIV positive (n=1188)** | 771 (98.6%) | 398 (98.0%) | 1,169 (98.4%) |
| **Body mass index (kg/m2)** |  |  |  |
| Underweight (<18.5) | 250 (5.5%) | 383 (11.7%) | 633 (8.1%) |
| Normal weight (18.5–24.9) | 1,678 (37.2%) | 2,165 (66.1%) | 3,843 (49.4%) |
| Overweight (25.0–29.9) | 1,323 (29.3%) | 548 (16.7%) | 1,871 (24.0%) |
| Obese (≥30) | 1,259 (27.9%) | 181 (5.5%) | 1,440 (18.5%) |
| Missing | 50 | 37 | 87 |
| **Abdominal circumference (cm), median (IQR)** | 86 (78, 97) | 80 (75, 87) | 83 (76, 93) |
| Missing | 400 | 368 | 768 |
| **Physical activity** |  |  |  |
| Low physical activity | 625 (13.8%) | 360 (11.0%) | 985 (12.6%) |
| Moderate physical activity | 701 (15.5%) | 345 (10.5%) | 1,046 (13.4%) |
| High physical activity | 3,201 (70.7%) | 2,575 (78.5%) | 5,776 (74.0%) |
| Missing | 33 | 34 | 67 |
| **Fruit intake (servings/day), median (IQR)<sup>1</sup>** | 0.14 (0.00, 0.86) | 0.14 (0.00, 0.86) | 0.14 (0.00, 0.86) |
| **Vegetable intake (servings/day), median (IQR)<sup>1</sup>** | 1.14 (0.57, 2.14) | 0.86 (0.29, 2.00) | 1.00 (0.43, 2.00) |
| **Consuming ≥5 portions of vegetables and fruits/day<sup>1</sup>** | 516 (11.3%) | 350 (10.6%) | 866 (11.0%) |
| **Always or often eating processed foods high in salt<sup>1</sup>** | 380 (8.3%) | 298 (9.0%) | 678 (8.6%) |
| **Always or often adding salt to food before or during eating<sup>1</sup>** | 631 (13.8%) | 523 (15.8%) | 1,154 (14.7%) |
| **Frequency of sweet food consumption<sup>1</sup>** |  |  |  |
| None per week | 2,143 (47.0%) | 1,821 (55.0%) | 3,964 (50.4%) |
| One to two times a week | 1,886 (41.4%) | 1,156 (34.9%) | 3,042 (38.6%) |
| Three to four times a week | 369 (8.1%) | 239 (7.2%) | 608 (7.7%) |
| Five or more times a week | 161 (3.5%) | 96 (2.9%) | 257 (3.3%) |
| **Frequency of sweet beverage consumption<sup>1</sup>** |  |  |  |
| None per week | 2,371 (52.0%) | 1,736 (52.4%) | 4,107 (52.2%) |
| One to two times a week | 1,762 (38.6%) | 1,271 (38.4%) | 3,033 (38.5%) |
| Three to four times a week | 291 (6.4%) | 203 (6.1%) | 494 (6.3%) |
| Five or more times a week | 135 (3.0%) | 102 (3.1%) | 237 (3.0%) |
| **Frequency of high-carbohydrate fried food consumption<sup>1</sup>** |  |  |  |
| None per week | 1,361 (29.9%) | 1,118 (33.8%) | 2,479 (31.5%) |
| One to two times a week | 2,215 (48.6%) | 1,585 (47.9%) | 3,800 (48.3%) |
| Three to four times a week | 706 (15.5%) | 410 (12.4%) | 1,116 (14.2%) |
| Five or more times a week | 277 (6.1%) | 199 (6.0%) | 476 (6.0%) |
| <sup>1</sup>Missing | 1 | 2 | 3 |
| **Smoking status** |  |  |  |
| Current Smoker | 1,015 (22.3%) | 1,579 (47.9%) | 2,594 (33.1%) |
| Never Smoker | 3,500 (77.0%) | 1,657 (50.2%) | 5,157 (65.8%) |
| Ex-smoker | 29 (0.7%) | 63 (1.9%) | 92 (1.2%) |
| Missing | 16 | 15 | 31 |
| **Frequency of alcohol consumption** |  |  |  |
| Every day/ 7 days per week | 71 (1.6%) | 158 (4.8%) | 229 (2.9%) |
| 5 to 6 days per week | 43 (0.9%) | 102 (3.1%) | 145 (1.8%) |
| 3 to 4 days per week | 93 (2.0%) | 183 (5.5%) | 276 (3.5%) |
| 1 to 2 days per week | 287 (6.3%) | 505 (15.3%) | 792 (10.1%) |
| Less than 1 day per week | 319 (7.1%) | 440 (13.3%) | 759 (9.7%) |
| Never | 3,729 (82.1%) | 1,914 (58.0%) | 5,643 (71.9%) |
| Missing | 18 | 12 | 30 |
| First-degree relative with hypertension | 976 (21.4%) | 374 (11.3%) | 1,350 (17.1%) |
| First-degree relative with diabetes | 257 (5.6%) | 90 (2.7%) | 347 (4.4%) |
| History of myocardial infarction | 62 (1.4%) | 11 (0.3%) | 73 (0.9%) |
| History of stroke | 20 (0.4%) | 13 (0.4%) | 33 (0.4%) |
| History of chronic kidney disease | 16 (0.4%) | 5 (0.2%) | 21 (0.3%) |
| Current or previous TB diagnosis | 169 (3.7%) | 238 (7.2%) | 407 (5.2%) |
| Severe vision impairment/blindness | 35 (0.8%) | 7 (0.2%) | 42 (0.5%) |
| Peripheral arterial disease | 27 (0.6%) | 5 (0.2%) | 32 (0.4%) |
| Heart failure | 32 (0.7%) | 5 (0.2%) | 37 (0.5%) |
| History of neuropathy | 39 (0.9%) | 8 (0.2%) | 47 (0.6%) |
| Chronic lung disease (COPD/asthma) | 16 (0.4%) | 11 (0.3%) | 27 (0.3%) |
| Diabetic foot syndrome | 6 (0.1%) | 1 (<0.1%) | 7 (0.1%) |

The self-reported HIV prevalence is 15.1%, with 98.4% of people reporting to live with HIV taking ART.

42.5% of adult participants are overweight (BMI 25 – 29.9 kg/m^2^) or obese (BMI ≥30 kg/m^2^). Obesity is more prevalent among women (27.9% versus 5.5%) while underweight (BMI <18.5 kg/m^2^) is more prevalent among men (11.7% versus 5.5%). Most participants (74.0%) report a high level of physical activity. Mean daily fruit and vegetable intake is 0.14 and 1.00 portions per day, respectively. 11% of participants meet the recommended minimal fruit and vegetable intake of five or more portions per day. 8.6% of participants always or often consume processed food high in salt and 14.7% always or often add salt to their food before or during eating. 33.1% of participants are smokers with higher rates among men (47.9% versus 22.3%). Overall, 71.9% of participants report to never consume alcohol (82.1% among women, 58% among men), while 2.9% drink daily (1.6% among women, 4.8% among men). 17.1% and 4.4% of participants have a first-degree relative living with hypertension or diabetes, respectively. 5.2% (7.2% male versus 3.7% female) report a current or previous tuberculosis infection while the self-reported prevalences or history of cardiovascular or diabetes complications (history of myocardial infarction, history of stroke, heart failure, peripheral arterial disease, chronic kidney disease, peripheral neuropathy, severe vision impairment/blindness, COPD or asthma, diabetic foot syndrome) range between 0.1% and 1%.

### Hypertension screening and care cascade

Of the 7’917 adult participants, for 7’763 (98%) arterial hypertension screening outcomes are available. Five participants report the intake of antihypertensive medication and are thus considered having hypertension but have missing blood pressure measurements. Table 4 shows the overview of the blood pressure screening outcomes and care cascade. The median systolic and diastolic blood pressures were 115 mmHg (IQR 106, 124) and 76 mmHg (IQR 70, 82) with similar values for women and men. 9.6% of women and 4.6% of men have a blood pressure of 140/90 mmHg or higher. The hypertension prevalence is 23.3% among women and 8.6% among men. Of the 1330 participants with hypertension, 1052 (79.1%) are women and 280 (21.05%) are men. 79.8% of participants with hypertension were aware of their diagnosis with higher awareness rates among women (82.8%) than men (68.3%). 75.1% of participants with hypertension are taking antihypertensive medication with higher treatment rates among women (78.3%) than men (62.9%). The control rate for hypertension is 56.1% with higher rates among women (58.5%) than men (46.8%).

**Table 4.** Overview of the hypertension screening outcomes and care cascade.

| Characteristic | Female, n (%),<br>4513 (58%) | Male, n (%),<br>3250 (42%) | Total, n (%),<br>7763 (100%) |
| --- | --- | --- | --- |
| **Systolic blood pressure<sup>1</sup> (mmHg), median (IQR)** | 114 (105, 125) | 116 (107, 124) | 115 (106, 124) |
| Missing | 5 | 0 | 5 |
| **Diastolic blood pressure<sup>1</sup> (mmHg), median (IQR)** | 76 (70, 83) | 75 (69, 81) | 76 (70, 82) |
| Missing | 5 | 0 | 5 |
| **Blood pressure categories** |  |  |  |
| Below 130/85 mmHg | 3,394 (75.2%) | 2,595 (79.9%) | 5,989 (77.2%) |
| 130-139/85-89 mmHg | 683 (15.1%) | 506 (15.6%) | 1,189 (15.3%) |
| Equal or above 140/90 mmHg | 434 (9.6%) | 148 (4.6%) | 582 (7.5%) |
| Missing | 5 | 0 | 5 |
| **Hypertension diagnosis status<sup>2</sup>** |  |  |  |
| Hypertensive | 1,052 (23.3%) | 278 (8.6%) | 1,330 (17.1%) |
| Not hypertensive | 3,464 (76.7%) | 2,971 (91.4%) | 6,435 (82.9%) |
| **Hypertension diagnosis awareness<sup>3</sup> (n=1330)** |  |  |  |
| Yes | 871 (82.8%) | 190 (68.3%) | 1,061 (79.8%) |
| No | 181 (17.2%) | 88 (31.7%) | 269 (20.2%) |
| **Receiving antihypertensive treatment (n=1330)** |  |  |  |
| Yes | 824 (78.3%) | 175 (62.9%) | 999 (75.1%) |
| No | 228 (21.7%) | 103 (37.1%) | 331 (24.9%) |
| **Controlled blood pressure<sup>4</sup> (n=1330)** |  |  |  |
| Yes | 613 (58.5%) | 130 (46.8%) | 743 (56.1%) |
| No | 434 (41.5%) | 148 (53.2%) | 582 (43.9%) |
| Missing | 5 | 0 | 5 |

<sup>1</sup> Calculated as the mean of the last two out of three consecutive measurements at intervals of one minute.
<sup>2</sup> Hypertension defined as two elevated blood pressure measurements in the range of 140-179/90-109 mmHg on two different days or two measurements of 180/110 mmHg or higher on the same day, at least 30 minutes apart or reporting intake of antihypertensive medication.
<sup>3</sup> Diagnosis awareness among people with hypertension defined as reporting to have been previously diagnosed with hypertension.
<sup>4</sup> Controlled blood pressure among people with hypertension defined as having a blood pressure below 140/90 mmHg.

### Diabetes screening and care cascade

5520 adult participants are 40 years or older or have a BMI of 25 kg/m^2^ or higher or reported intake of antidiabetic medication and are therefore eligible for diabetes screening. Diabetes screening and care cascade outcomes were available for 5428 participants (98% of those eligible for diabetes screening) and are shown in table 5. Median fasting and random blood glucose levels were 4.6 and 4.9 mmol/l. 5.2% of women and 2.0% of men have prediabetes. Type 2 diabetes prevalence is 4.0% with higher prevalence among women (4.8%) than men (2.6%). Two women have type 1 diabetes. Of the 220 participants with type 2 diabetes, 149 (67.1%) reported current use of antidiabetic medication. 38.8% of participants with type 2 diabetes had controlled blood glucose levels. Treatment coverage was the same among men and women, while women had a better control rate (42.4% versus 26.5%). Median fasting blood glucose among people with type 2 diabetes was 7.8 mmol/l (4 (1.86%) missing) and median HbA1c was 6.7% (25 (12.9%) missing).

**Table 5.** Overview of the diabetes screening outcomes and care cascade of participants with type 2 diabetes. HbA1c: Glycated haemoglobin, IQR: interquartile range.

| Characteristic | Female, n (%),<br>3507 (65%) | Male, n (%),<br>1921 (35%) | Total, n (%),<br>5428 (100%) |
| --- | --- | --- | --- |
| **Fasting blood glucose (mmol/l), median (IQR)** | 4.90 (4.40, 5.30) | 4.60 (4.10, 5.10) | 4.80 (4.30, 5.20) |
| Missing | 588 | 336 | 924 |
| **Random blood glucose (mmol/l), median (IQR)** | 5.00 (4.40, 5.30) | 4.80 (4.30, 5.30) | 4.90 (4.40, 5.30) |
| Missing | 2,868 | 1,574 | 4,442 |
| **Diabetes diagnosis status<sup>1</sup>** |  |  |  |
| Screened normal | 3,151 (89.9%) | 1,832 (95.5%) | 4,983 (91.9%) |
| Prediabetes prevalence | 183 (5.2%) | 38 (2.0%) | 221 (4.1%) |
| Type 1 diabetes prevalence | 2 (<0.1%) | 0 (0%) | 2 (<0.1%) |
| Type 2 diabetes prevalence | 170 (4.8%) | 49 (2.6%) | 219 (4.0%) |
| **Diabetes diagnosis awareness (n=219)<sup>2</sup>** |  |  |  |
| Yes | 110 (64.7%) | 33 (67.3%) | 147 (67.1%) |
| No | 60 (35.3%) | 16 (32.7%) | 76 (34.7%) |
| **Current use of diabetes treatment (n=220)** |  |  |  |
| Yes | 114 (67.1%) | 33 (67.3%) | 147 (67.1%) |
| No | 56 (32.9%) | 16 (32.7%) | 72 (32.9%) |
| **Controlled diabetes (n=220)<sup>3</sup>** |  |  |  |
| Yes | 72 (42.4%) | 13 (26.5%) | 85 (38.8%) |
| No | 98 (57.6%) | 36 (73.5%) | 134 (61.2%) |
| **Fasting blood glucose (mmol/l) (n=215), median (IQR)** | 7.5 (5.7, 9.9) | 8.7 (6.6, 9.9) | 7.8 (6.0, 9.9) |
| Missing | 4 | 0 | 4 |
| **Random blood glucose (mmol/l) (n=63), median (IQR)** | 11.9 (7.8, 16.5) | 9.9 (8.3, 11.5) | 11.3 (8.0, 16.4) |
| Missing | 117 | 39 | 156 |
| **HbA1c (%) (n=194), median (IQR)** | 6.60 (5.80, 8.50) | 7.00 (6.10, 7.60) | 6.70 (5.80, 8.40) |
| Missing | 21 | 4 | 25 |

<sup>1</sup> Diabetes defined as random blood sugar $\geq 11.1$ mmol/l or fasting blood sugar $\geq 7.0$ mmol/l either in presence of cardinal diabetes symptoms (polyuria, polydipsia and weight loss) or after previously elevated blood sugar on a different day or reporting intake of antidiabetic medication. Classification as type 1 or type 2 based on previous health records and clinical expert opinion considering age at diagnosis, BMI, and family history.
<sup>2</sup> Diagnosis awareness defined as reporting to have been previously diagnosed with diabetes.
<sup>3</sup> Controlled diabetes defined as having a fasting blood sugar $< 7.0$ mmol/l or a HbA1c $< 6.5\%$ if fasting blood glucose is missing.

## Findings to date

### Socioeconomic and demographic characteristics

The ComBaCaL cohort reflects the socioeconomic realities of rural Lesotho, characterized by limited economic resources and poor access to essential infrastructure. The IWI score of 26 indicates poor quality of housing material and scarcity of durable assets [51] and the median monetary monthly household income of 42.4 USD is far below the international poverty line of 2.15 USD per person per day, which defines extreme poverty [63]. Both the IWI score as well as the monetary income in our population are below the Lesotho national average, underscoring the economic disadvantages in our rural study area compared to the general Lesotho population [64,65].

These socioeconomic conditions have significant public health implications through various pathways. Access to affordable, reliable and clean energy is essential for health and development [66,67]. However, only 13.2% of ComBaCaL cohort households have access to electricity, with the majority relying on polluting fuels like biomass and paraffin that have well-known negative effects on lung health, cardiovascular risk and other health outcomes [68,69]. Additionally, poor sanitation is a major concern with most households using unimproved pit latrines and nearly 40% of households lacking any toilet facilities. Poor sanitation contributes to the transmission of gastrointestinal infections, a leading cause of death among young children globally and a significantly contributor to Lesotho’s high under-5 mortality rate of 72 deaths 1’000 live births [70,71].

In line with a recent assessment of food insecurity among small-holder farming households in Lesotho [72], 49% of ComBaCaL cohort households experience moderate or severe food insecurity. Food insecurity reduces life expectancy not only through malnourishment, but also by contributing to the development of non-communicable diseases [73,74]. Although most households in the cohort own agricultural land, this does not seem to offer effective protection from food insecurity, due to the low levels of mechanisation and commercialization of small-holder farming in our region, which limits economic development and creates vulnerabilities to weather shocks and other environmental or economic challenges [75,76]. In July 2024, the Lesotho Prime Minister declared a state of emergency due to a severe food crisis caused by a prolonged drought, calling for international support to improving nutrition through enhanced agricultural production and resilience [77].

The limited economic resources, coupled with geographically challenging terrain and poor transport infrastructure, impede access to essential healthcare services and medications. This underscores the need for more accessible and equitable healthcare delivery systems for the rural areas, such as community-based health service models.

With a median age of 19 years, our cohort is younger than the general Lesotho population (median age 22 years), while the proportion of elderly individuals (9.2% versus 6.1%) and women (55% versus 51%) is higher than at the national level [29,78]. These demographic patterns may be linked to labour migration, which has been a dominating feature of the Lesotho economy, with people of working age, predominantly men, leaving the rural areas for mine work or other jobs in South Africa or urban areas of Lesotho [79–81]. Furthermore, the higher fertility rates in rural compared to urban areas may contribute to the younger median age in our cohort [82,83].

The cohort’s educational attainment is significantly lower than the national average where the most recent Demographic Health Survey reported 75% of women and 59% of men to have at least secondary school education [83]. This disparity underscores the educational disadvantages faced by rural populations in Lesotho with implications on economic development, health literacy, and thus health outcomes [84].

The formal employment rate is low (13%) while self-employment (18%) and agriculture (20%) are the most prevalent work statuses. 15% of participants reported to be unemployed, which is slightly below the national average of 16.5% [85].

### Chronic disease risk factors and complications

In line with a recent population-based survey conducted in the same region [25] and the national estimates [86], we observe a considerable rate of overweight (29.3%) and obesity (27.9%) among women, while men mostly have normal weight (66.1%) with lower but still relevant rates of overweight (16.7%) and obesity (5.5%). Like in many other low-and-middle-income countries, the rates of obesity in Lesotho have increased considerably over the last decades with an estimated three-fold increase since 1990 [86]. The high rates of overweight and obesity in our cohort confirm that the increasing obesity prevalence does not only affect the urban population but has a similar impact in rural areas. Importantly, underweight, especially among men, still poses a significant health burden in our cohort, a pattern observed in similar African settings [86].

A high level of physical activity was reported for men (78.5%) and women (70.7%), while the reported consumption of unhealthy food items and salt was low. Despite the limited correlation of self-reported dietary and physical activity patterns with objective measures [87–89], it appears that physical inactivity and consumption of typical unhealthy food items are not the main drivers for overweight in our rural study population. Other dietary factors, such as the overconsumption of widely available plain starch, that were not captured by our assessments might be contributing to the high rates of overweight and further investigations of dietary patterns are needed to fully understand the dietary factors contributing to the high rates of abnormal weight.

Fruit and vegetable consumption in our study population is low, with only 11% reaching the WHO-recommended minimum intake of five portions per day [90]. This rate is lower than in any country included in a recent systematic review on fruit and vegetable consumption in low-and-middle-income countries and part of the above-mentioned food insecurity crisis [91]. The particularly low fruit consumption with an average of only 0.14 portions per day is likely to be attributed to the low availability and relatively high prices of fruits in the region. Lesotho is a high-altitude country, and small-scale crop and cattle production are the main agricultural outputs while local fruit production is minimal [92]. Low fruit consumption is the leading dietary risk factor in terms of disability-adjusted life years and deaths in Southern Africa [93] and the low fruit and vegetable consumption has been identified as main gap towards a healthy and diverse diet for Basotho households [92]. To reduce diet-related health burden, efforts are urgently needed to transform agricultural practices and develop more resilient food supply systems, thereby improving food security including access to a diverse diet with sufficient fresh fruits and vegetables [72,94].

The smoking rates of 47.9% among men and 22.3% among women are higher than reported in a recent population-based survey [25] and also higher than the global average [95]. In Lesotho, smoking rates among men have remained stable on a high level while female smoking rates have increased sharply over the last decade [96] – a concerning pattern observed in many low- and middle-income countries [97].

Alcohol use at least once a week was reported by 18.2% of participants with higher frequencies for men, slightly exceeding the national estimates [98]. As in many other low-and-middle-income countries, alcohol consumption in Lesotho has increased considerably over the last two decades, a trend particularly concerning given the increased vulnerability of poor people to the deleterious effect of alcohol [98,99]. While alcohol consumption at the current level may contribute less to the overall NCD burden in the ComBaCaL cohort compared to other risk factors, such as smoking or obesity, it remains a concern as it is a risk factor for the development of NCDs [100], and is known to increase the transmission of HIV and the susceptibility to other infectious diseases, especially tuberculosis [98].

Lesotho is among the countries with the highest HIV and tuberculosis burden globally [30,101] and the communicable disease burden currently remains higher than the NCD-related burden [100]. This is reflected in our cohort, where the self-reported HIV prevalence is 15.1%, the reported lifetime tuberculosis prevalence is 5.2%, while prevalences of cardiovascular disease complications are 1% or less. In the most recent population-based HIV impact assessment, the HIV prevalence assessed through population-based HIV testing in Mokhotlong and Butha-Buthe districts was 19% [26]. The comparably lower self-reported HIV prevalence in our cohort could be attributed to lower infection rates in the rural compared to urban areas, underreporting by participants or unawareness of diagnosis.

The low rates of cardiovascular disease complications reported in our cohort warrant further investigation. First, the diagnostic options for cardiovascular disease complications available in Lesotho are limited, leading to potential underdiagnosis. Second, due to limited therapeutic options and a generally low life expectancy [102], age-related cardiovascular disease complications may be less prevalent compared to other settings. A recent study conducted in Butha-Buthe and Mokhotlong has revealed high rates of undiagnosed end-organ damage associated with cardiovascular diseases, such as renal impairment, left-ventricular remodelling and peripheral neuropathy [32]. Longitudinal observations including cause of death assessments and more extensive diagnostics are required to provide further context to the available self-reported information. A recent echocardiography study in Butha-Buthe has revealed high levels of chronic pulmonary heart disease [103] and considering the high smoking rates, the high tuberculosis prevalence and occupational risk exposure of mine work, it seems likely that the true prevalence of chronic lung disease is higher than the 0.3% reported by participants in this study.

### Hypertension and diabetes care cascades

The hypertension prevalence of 17% (23% among women, 8.6% among men) is slightly lower than reported in a recent population-based survey in the two districts (22%) [25] and considerably lower than reported by a WHO survey at national level in 2012 (31.0%) [96]. These differences could be attributed to lower prevalences in rural compared to urban areas and potential overreporting in surveys that rely on single-day blood pressure measurements. Globally, the hypertension prevalence is slightly higher among men than women [104]. The higher prevalence among women in our cohort is likely to be driven by the higher overweight and obesity rates among women [105]. However, gender-specific age structures with more older females and differing care-seeking behaviours, with higher rates of previous diagnosis among women could also contribute to the observed gender disparity [106]. The diagnosis awareness rate (80%), treatment rate (75%) and control rate (56%) are considerably higher than in other low-and-middle-income countries, including Southern African countries [107,108]. This finding is remarkable, especially considering the high proportion of participants living in remote areas with difficult access to healthcare services and potentially lower level of health literacy due to limited formal education. The higher awareness, treatment and control rates in women compared to men are likely to be attributed to differences in healthcare seeking behaviour. It remains to be investigated whether the comparably high awareness, treatment and control rates could be influenced by potential overdiagnosis and overtreatment. Despite these findings, a substantial care gap remains with almost half of people living with hypertension not reaching treatment targets.

The type 2 diabetes prevalence in our cohort of 4.0% (4.8% among women, 2.5% among men) is lower than in a recent population-based survey in the two districts (5.3%) [25] and in the WHO survey conducted in 2012 (6.3%) [96]. This could be attributed to a potentially lower prevalence in rural settings and to the use of different diagnostic criteria. In our cohort we took blood sugar measurements on different days while other studies often rely on single-day measurements. The observed treatment and control rates (67% and 39%) are higher than in most low-and-middle income countries and much higher than in the neighbouring country South Africa, where only 8.7% of people living with diabetes were reported to reach adequate control despite the use of a less stringent target level (8 mmol/l versus 7 mmol/l) [109,110]. The remaining care gap remains substantial with 61% of people living with diabetes not reaching treatment targets. The possibility of overdiagnosis and overtreatment also warrants further investigation.

### Strengths and limitations

Our study provides a comprehensive overview of sociodemographic characteristics, chronic disease risk factors, cardiovascular complications, HIV prevalence and on the care cascades for hypertension and diabetes in rural Lesotho. We demonstrate that trained CHWs, supported by a suitable clinical decision data collection tool, can facilitate large-scale population-based research data collection. Data collection by CHWs who live in the study villages offers the possibility of repeated visits allowing for very high enrolment, screening and follow-up rates. Continuous remote monitoring and interactions with CHWs through phone calls or field visits ensures high data accuracy and completeness. All CHWs in the ComBaCaL cohort operate within the Lesotho Ministry of Health framework.

The use of the TwiCs design enables the efficient implementation of trials nested in the cohort to generate evidence on healthcare interventions at a large-scale.

Limitations include the self-reported nature of many outcomes with biomedical assessments only conducted for hypertension and diabetes. Besides the inherent shortcomings of self-reported behavioural outcomes, the reliability of self-reported chronic disease complications is limited by the insufficient availability of appropriate diagnostics in Lesotho and by the limited clinical expertise of lay CHWs. This is partially mitigated by the careful training of CHWs on reading medical notes in the participant’s health booklets and verification of reported diagnoses by the study team remotely or through field visits. For the assessment of food insecurity, only six out of the nine questions of the HFIAS were included in our questionnaire leading to limited sensitivity and making the calculation of the HFIAS score impossible [53]. People reporting intake of antihypertensive, or antidiabetic medication were reported as living with the respective condition without further verification of the diagnosis made by a previous healthcare provider. Thus, potential overdiagnosis and overtreatment cannot be excluded. To date, no HIV tests have been conducted and the associations between HIV and NCDs, as well as the interplay of shared risk factors have not yet been explored in detail. Furthermore, no anthropometric and behavioural risk factor data was collected among children impeding investigations of early chronic disease risk factors.

### Future plans

Currently, three trials nested within the cohort assessing the effectiveness of CHW-delivered care for hypertension and diabetes are ongoing and will be completed early 2025. The prospective follow-up of participants living with hypertension and diabetes as part of the nested trials will also allow for a closer investigation of potential overdiagnosis and overtreatment of these conditions.

Further follow-up of the cohort including assessments of causes of death will allow for a more comprehensive understanding of the burden of different disease entities. The prevalence of behavioural risk factors including smoking, alcohol consumption, dietary and physical activity patterns will be assessed prospectively as part of future cohort follow-up visits which to generate information on the dynamics of these risk factors over time.

The next trial to be nested within the cohort is in preparation and will assess the effectiveness of CHW-delivered HIV prevention and care. As part of this, HIV testing will be added to the cohort assessments, which will provide further insights into the true HIV prevalence and allow for more detailed investigations of associations between behavioural risk factors, NCDs and HIV. Furthermore, this trial will generate evidence on the feasibility and effectiveness of integrated CHW-delivered care targeting HIV, diabetes and hypertension in a comprehensive community-based care package.

## Conclusion

The ComBaCaL cohort is a representative sample of the population living in rural Lesotho. It highlights significant socioeconomic challenges, such as low levels of wealth, education, housing quality, sanitation, access to clean energy, secure and diverse food supply, and health services compared to global and national standards, thereby posing multiple health risks. In addition to the anticipated high prevalence of HIV, we observed significant rates of hypertension and diabetes. While treatment and control rates for these chronic conditions are higher than expected, substantial care gaps remain. The most important behavioural risk factors identified are the high rates of overweight and obesity among women and the high smoking rates among men.

Beyond providing detailed socioeconomic, behavioural, and clinical insights, the ComBaCaL cohort, through its nested trials using the Trials within Cohorts (TwiCs) design, will generate evidence on the effectiveness of Community Health Worker (CHW)-delivered interventions to improve access to quality care for diabetes, hypertension, and HIV. The findings from the ComBaCaL cohort and its nested trials will enhance the understanding of chronic disease dynamics and inform the development of targeted health interventions in Lesotho and other regions with similar resource-limited, rural characteristics, particularly those employing a similar CHW system.

## Data Availability

De-identified data will be made available on a suitable repository, such as zenodo or dryad. Investigators who would like to use the data for scientific publications must submit a concept sheet detailing the required data and planned analyses to the corresponding author for internal review and approval.

## Further details

## Ethics approval

The ComBaCaL cohort study and ongoing nested TwiCs were approved by the National Health Research Ethics Committee in Lesotho (ID 210-2022 and ID 210-2022 Nested) and the Ethikkommission Nordwest- und Zentralschweiz in Switzerland (AO_2022-058, AO_2022-00074 and AO_2022-00077) and are registered on clinicaltrials.gov (NCT05596773, NCT05743387, and NCT05684055). Amendments to the cohort protocol will be submitted to the National Health Research Ethics Committee in Lesotho prior to implementation.

## Patient and public involvement

The Ministry of Health of Lesotho and the District Management Teams of Butha-Buthe district were among the initial members of the research consortium. Community-Engaged Research is key in the ComBaCaL program. The CHWs are members of the community and have been elected by the community. Research questions for nested trials are based on feedback from community-members during implementation of the ComBaCaL activities. Prior to implementation of the ComBaCaL cohort, we conducted a pilot study to incorporate views and perceptions of the local community and CHWs in the final design of the study and nested trials. Community representatives will be involved in the development of protocols for further nested trials.

## Collaboration and Data Sharing

De-identified data will be made available on a suitable repository, such as zenodo or dryad. Investigators who would like to use the data for scientific publications must submit a concept sheet detailing the required data and planned analyses to the corresponding author for internal review and approval. ComBaCaL investigators or contributors shall be co-authors on publications using ComBaCaL data, provided they fulfil authorship criteria as defined by the International Committee of Medical Journal Editors [111].

## Author contributions

AA and NDL are the principal investigators, they acquired the main funding, lead the project and substantially contributed to the manuscript. FG drafted the manuscript, led the clinical development of the ComBaCaL app, the study implementation, and conceptualized the study together with the principal investigators. PG was responsible for the central management of the study implementation and contributed to the conceptualization of the study. RG (lead), TIL, MC, TK, MalebM, MalehM, MosM, MotM, ManM, MK, PMS, MB and RM were responsible for the local implementation of the study through training and supervision of CHWs and local data monitoring. TT, and FR supported the development of the ComBaCaL app and the training of VHWs. TL and GSS were responsible for central data management and GSS contributed substantially to the manuscript writing. SM was responsible for the local data management. MadM, SP, LS, MosT and MalL are responsible for the collaboration with the Lesotho Ministry of Health and gave input on the study design to ensure alignment with local guidelines and practices. DB and KK led the technical development of the ComBaCaL app. TB and ES are clinical hypertension and diabetes experts. They reviewed and approved the clinical algorithms and gave input on the presentation of the outcomes. MB is a clinical trial methods expert and gave input on the study design. FC is responsible for statistical analysis. All authors read and approved the final manuscript.

## Acknowledgements

We would like to acknowledge the SolidarMed team in Lesotho and the involved Community Health Workers (CHWs) for their essential contributions to the ComBaCaL project. Our thanks also go to Ilse van Roy from SolidarMed Switzerland for her contributions as part of the project steering committee. We are grateful to the team at Medic (www.medic.org) for making the Community Health Toolkit openly available and for the technical support provided. Additionally, we extend our acknowledgements to the students from the Department of Informatics at the University of Zurich, under the supervision of Dario Staehelin and Gerhard Schwabe, for their contributions to developing the pilot version of the ComBaCaL app.

