## Supplemental Tables for "Cohort profile: design, sociodemographic characteristics, chronic disease risk factors, and baseline hypertension and diabetes care cascades of the open, prospective Community-Based chronic disease Care Lesotho (ComBaCaL) cohort"

| Characteristic | N = 5,008 |
| --- | --- |
| **District** | |
| Butha-Buthe | 2,486 (49.6%) |
| Mokhotlong | 2,522 (50.4%) |
| **Number of household members, median (IQR)** | 3 (2, 4) |
| **International Wealth Index, median (IQR)** | 26 (17, 36) |
| **Monthly household income (USD), median (IQR)[1]** | 42.4 (18.6, 63.6) |
| Unknown | 7 |
| **Main source of household income** | |
| Salary/wages[2] | 1,289 (25.7%) |
| Informal business (e.g. street vendor) | 1,109 (22.1%) |
| Formal business (e.g. retail trade) | 37 (0.7%) |
| Farming | 1,213 (24.2%) |
| Rentals / interest | 61 (1.2%) |
| Grants / pension | 1,109 (22.1%) |
| Refused to say | 190 (3.8%) |
| **Access to health facility** | |
| Easy access | 2,460 (49.1%) |
| Hard access[3] | 2,548 (50.9%) |
| **Difficulties accessing health care[4]** | 1,120 (22.4%) |
| **Difficulties accessing medications[5]** | 821 (16.4%) |
| **Household Food Insecurity Access category** | |
| Food secure | 2,385 (47.6%) |
| Mildly food insecure | 167 (3.3%) |
| Moderately food insecure | 965 (19.3%) |
| Severely food insecure | 1,489 (29.7%) |
| Unknown | 2 |
| **Drinking water** | |
| Running (piped) water inside the dwelling | 15 (0.3%) |
| Running (piped) water outside dwelling but inside yard | 64 (1.3%) |
| Neighbor’s tap | 43 (0.9%) |
| Public / community tap | 4,075 (81.4%) |
| Borehole | 548 (10.9%) |
| Dam/ river | 216 (4.3%) |
| Rainwater tank | 41 (0.8%) |
| Bottled water | 0 (0.0%) |
| Refused to say | 6 (0.1%) |
| **Toilet** | |
| Own flush toilet | 21 (0.4%) |
| Flush toilet shared with another household | 12 (0.2%) |
| Own ventilated improved pit latrine | 628 (12.5%) |
| Own pit latrine with slab | 1,993 (39.8%) |

| Characteristic | N = 5,008 |
|---|---|
| Open pit latrine | 179 (3.6%) |
| Composting toilet | 68 (1.4%) |
| None/Bush/Field | 1,959 (39.1%) |
| Pit latrine shared with another household | 141 (2.8%) |
| Refused to say | 7 (0.1%) |
| **Electricity available** | 663 (13.2%) |
| **Cooking fuel** | |
| Electricity | 106 (2.1%) |
| LPG | 208 (4.2%) |
| Biogas | 29 (0.6%) |
| Paraffin | 147 (2.9%) |
| Wood | 4,210 (84.1%) |
| Coal | 4 (0.1%) |
| Straw, shrubs, grass | 16 (0.3%) |
| Animal dung | 286 (5.7%) |
| Refused to say | 2 (0.0%) |
| **Heating fuel** | |
| Electricity | 68 (1.4%) |
| Gas | 41 (0.8%) |
| Paraffin | 323 (6.4%) |
| Wood | 3,198 (63.9%) |
| Coal | 23 (0.5%) |
| Animal dung | 1,308 (26.1%) |
| Crop waste | 13 (0.3%) |
| Refused to say | 34 (0.7%) |
| **People per bedroom** | |
| 1-2 | 2,527 (50.5%) |
| 3-4 | 1,880 (37.5%) |
| 5 or more | 599 (12.0%) |
| Other / unknown / refused to say | 2 (0.0%) |
| **Wall material** | |
| Mud | 3,099 (61.9%) |
| Mud and cement | 1,042 (20.8%) |
| Corrugated iron | 230 (4.6%) |
| Bare brick | 245 (4.9%) |
| Finished / plaster | 348 (6.9%) |
| Other | 42 (0.8%) |
| Refused to say | 2 (0.0%) |
| **Roof material** | |
| Natural | 3,099 (61.9%) |
| Rudimentary (wood planks, cardboard) | 242 (4.8%) |

| Characteristic | N = 5,008 |
|---|---|
| Finished (metal, corrugated, wood, asbestos, ceramic / clay tiles, roofing shingles) | 1,548 (30.9%) |
| Other | 116 (2.3%) |
| Refused to say | 3 (0.1%) |
| **Floor material** | |
| Earth | 2,511 (50.1%) |
| Wood planks | 30 (0.6%) |
| Cement | 1,462 (29.2%) |
| Vinyl tiles | 203 (4.1%) |
| Ceramic tiles | 232 (4.6%) |
| Carpet | 549 (11.0%) |
| Other | 19 (0.4%) |
| Refused to say | 2 (0.0%) |
| **Agricultural land ownership** | 3,127 (62.4%) |
| **Clock** | 1,059 (21.1%) |
| **Water pump** | 83 (1.7%) |
| **Grain grinder** | 700 (14.0%) |
| **Sewing machine** | 1,093 (21.8%) |
| **Color TV** | 1,827 (36.5%) |
| **Black/white TV** | 1,620 (32.3%) |
| **Video player / DVD player** | 212 (4.2%) |
| **Tape player / CD player** | 150 (3.0%) |
| **Camera, video camera** | 39 (0.8%) |
| **Refrigerator** | 259 (5.2%) |
| **Freezer** | 66 (1.3%) |
| **Washing machine** | 12 (0.2%) |
| **Dishwasher** | 7 (0.1%) |
| **Electric or gas stove** | 1,063 (21.2%) |
| **Kerosene stove** | 1,878 (37.5%) |
| **Mobile phone** | 2,767 (55.3%) |
| **Car** | 135 (2.7%) |
| **Bicycle** | 26 (0.5%) |
| **Landline or fix phone** | 21 (0.4%) |
| **Cattle** | 0 (0, 2) |
| **Horses** | 0 (0, 0) |
| **Donkeys** | 0 (0, 1) |
| **Sheep** | 0 (0, 7) |
| **Goats** | 0 (0, 0) |
| **Chickens** | 0 (0, 5) |
| **Pigs** | 0 (0, 0) |
| **Rabbits** | 0 (0, 0) |

| Characteristic | N = 5,008 |
|---|---|

[1] Collected in Lesotho Loti, converted using the bilateral conversion rate on May 31, 2024 [62]
[2] Salaries and wages of household members or other people (i.e. of family members living outside the household)
[3] Defined as needing to cross a mountain or river or travel >10 km to the nearest health facility.
[4] Defined as at least one household member not being able to consult a healthcare provider despite needing it during the last 12 months.
[5] Defined as at least one household member experiencing difficulties obtaining required medication.

*Supplementary table 1 Overview of all collected household level socioeconomic indicators. IQR: interquartile range*

| Characteristic | Female, n(%), 4560 (58%) | Male, n(%), 3314 (42%) | Total, n(%), 7874 (100%) |
|---|---|---|---|
| **Age (years), median (IQR)** | 41 (27, 60) | 39 (28, 54) | 40 (27, 58) |
| 18-39 years | 2,148 (47.1%) | 1,662 (50.2%) | 3,810 (48.4%) |
| 40-64 years | 1,529 (33.5%) | 1,185 (35.8%) | 2,714 (34.5%) |
| ≥65 years | 883 (19.4%) | 467 (14.1%) | 1,350 (17.1%) |
| **District** | | | |
| Butha-Buthe | 2,283 (50.1%) | 1,592 (48.0%) | 3,875 (49.2%) |
| Mokhotlong | 2,277 (49.9%) | 1,722 (52.0%) | 3,999 (50.8%) |
| **Relationship status** | | | |
| Single | 655 (14.4%) | 902 (27.2%) | 1,557 (19.8%) |
| In a committed relationship | 2,690 (59.0%) | 2,024 (61.1%) | 4,714 (59.9%) |
| Separated or divorced | 87 (1.9%) | 123 (3.7%) | 210 (2.7%) |
| Widowed | 1,119 (24.5%) | 245 (7.4%) | 1,364 (17.3%) |
| Unknown/refused to say | 9 (0.2%) | 20 (0.6%) | 29 (0.4%) |
| **Education** | | | |
| No schooling | 276 (6.1%) | 684 (20.7%) | 960 (12.2%) |
| Primary school | 2,623 (57.5%) | 1,879 (56.7%) | 4,502 (57.2%) |
| Secondary/high school | 1,448 (31.8%) | 624 (18.8%) | 2,072 (26.3%) |
| Tertiary/higher/post high school | 209 (4.6%) | 121 (3.7%) | 330 (4.2%) |
| Unknown/refused to say | 3 (0.1%) | 4 (0.1%) | 7 (0.1%) |
| Missing | 1 | 2 | 3 |
| **Work status** | | | |
| Working for pay | 444 (9.7%) | 572 (17.3%) | 1,016 (12.9%) |
| Self-employed | 703 (15.4%) | 709 (21.4%) | 1,412 (17.9%) |
| Working on own plot or looking after livestock | 469 (10.3%) | 1,075 (32.4%) | 1,544 (19.6%) |
| Helping another family member with their business, without pay | 33 (0.7%) | 30 (0.9%) | 63 (0.8%) |
| Full-time student | 90 (2.0%) | 38 (1.1%) | 128 (1.6%) |
| Homemaker (looking after children/others/home) | 1,734 (38.0%) | 184 (5.6%) | 1,918 (24.4%) |
| Long term sick or disabled | 53 (1.2%) | 41 (1.2%) | 94 (1.2%) |
| Retired | 205 (4.5%) | 136 (4.1%) | 341 (4.3%) |
| Unemployed | 702 (15.4%) | 461 (13.9%) | 1,163 (14.8%) |
| Unknown/refused to say | 127 (2.8%) | 68 (2.1%) | 195 (2.5%) |
| **Body mass index (kg/m2)** | | | |
| Underweight (<18.5) | 250 (5.5%) | 383 (11.7%) | 633 (8.1%) |
| Normal weight (18.5–24.9) | 1,678 (37.2%) | 2,165 (66.1%) | 3,843 (49.4%) |
| Overweight (25.0-29.9) | 1,323 (29.3%) | 548 (16.7%) | 1,871 (24.0%) |
| Obese (≥30) | 1,259 (27.9%) | 181 (5.5%) | 1,440 (18.5%) |
| Missing | 50 | 37 | 87 |
| **Abdominal circumference (cm)** | 86 (78, 97) | 80 (75, 87) | 83 (76, 93) |
| Missing | 400 | 368 | 768 |

| Characteristic | Female, n(%), 4560 (58%) | Male, n(%), 3314 (42%) | Total, n(%), 7874 (100%) |
|---|---|---|---|
| **Physical activity** | | | |
| Low physical activity | 625 (13.8%) | 360 (11.0%) | 985 (12.6%) |
| Moderate physical activity | 701 (15.5%) | 345 (10.5%) | 1,046 (13.4%) |
| High physical activity | 3,201 (70.7%) | 2,575 (78.5%) | 5,776 (74.0%) |
| Missing | 33 | 34 | 67 |
| **Fruit intake (servings/day)** | 0.14 (0.00, 0.86) | 0.14 (0.00, 0.86) | 0.14 (0.00, 0.86) |
| Missing | 1 | 2 | 3 |
| **Vegetable intake (servings/day)** | 1.14 (0.57, 2.14) | 0.86 (0.29, 2.00) | 1.00 (0.43, 2.00) |
| Missing | 1 | 2 | 3 |
| **Portions of vegetables and fruits/day** | | | |
| <5 | 4,043 (88.7%) | 2,962 (89.4%) | 7,005 (89.0%) |
| ≥5 | 516 (11.3%) | 350 (10.6%) | 866 (11.0%) |
| Missing | 1 | 2 | 3 |
| **Adding salt during cooking** | | | |
| Always | 3,008 (66.0%) | 2,226 (67.2%) | 5,234 (66.5%) |
| Often | 130 (2.9%) | 123 (3.7%) | 253 (3.2%) |
| Sometimes | 773 (17.0%) | 523 (15.8%) | 1,296 (16.5%) |
| Rarely | 509 (11.2%) | 360 (10.9%) | 869 (11.0%) |
| Never | 131 (2.9%) | 68 (2.1%) | 199 (2.5%) |
| Unknown/refused to say | 8 (0.2%) | 12 (0.4%) | 20 (0.3%) |
| Missing | 1 | 2 | 3 |
| **Adding salt while eating** | | | |
| Always | 631 (13.8%) | 523 (15.8%) | 1,154 (14.7%) |
| Often | 206 (4.5%) | 219 (6.6%) | 425 (5.4%) |
| Sometimes | 1,295 (28.4%) | 1,028 (31.0%) | 2,323 (29.5%) |
| Rarely | 1,246 (27.3%) | 1,011 (30.5%) | 2,257 (28.7%) |
| Never | 1,175 (25.8%) | 521 (15.7%) | 1,696 (21.5%) |
| Unknown/refused to say | 6 (0.1%) | 10 (0.3%) | 16 (0.2%) |
| Missing | 1 | 2 | 3 |
| **Importance of lowering salt intake** | | | |
| Very important | 4,000 (87.7%) | 2,697 (81.4%) | 6,697 (85.1%) |
| Somewhat important | 145 (3.2%) | 164 (5.0%) | 309 (3.9%) |
| Not at all important | 110 (2.4%) | 133 (4.0%) | 243 (3.1%) |
| Unknown/refused to say | 304 (6.7%) | 318 (9.6%) | 622 (7.9%) |
| Missing | 1 | 2 | 3 |
| **Perceiving high salt intake as potential health problem** | | | |
| Yes | 3,775 (82.8%) | 2,504 (75.6%) | 6,279 (79.8%) |
| No | 519 (11.4%) | 502 (15.2%) | 1,021 (13.0%) |
| Unknown/refused to say | 265 (5.8%) | 306 (9.2%) | 571 (7.3%) |
| Missing | 1 | 2 | 3 |

| Characteristic | Female, n(%), 4560 (58%) | Male, n(%), 3314 (42%) | Total, n(%), 7874 (100%) |
|---|---|---|---|
| **Eating processed food high in salt** | | | |
| Always | 380 (8.3%) | 298 (9.0%) | 678 (8.6%) |
| Often | 217 (4.8%) | 223 (6.7%) | 440 (5.6%) |
| Sometimes | 1,460 (32.0%) | 1,048 (31.6%) | 2,508 (31.9%) |
| Rarely | 1,921 (42.1%) | 1,383 (41.8%) | 3,304 (42.0%) |
| Never | 560 (12.3%) | 346 (10.4%) | 906 (11.5%) |
| Unknown/refused to say | 21 (0.5%) | 14 (0.4%) | 35 (0.4%) |
| Missing | 1 | 2 | 3 |
| **Perceived quantity of salt intake** | | | |
| Far too much | 86 (1.9%) | 78 (2.4%) | 164 (2.1%) |
| Too much | 193 (4.2%) | 186 (5.6%) | 379 (4.8%) |
| Just the right amount | 2,879 (63.1%) | 2,108 (63.6%) | 4,987 (63.4%) |
| Too little | 1,177 (25.8%) | 786 (23.7%) | 1,963 (24.9%) |
| Far too little | 140 (3.1%) | 63 (1.9%) | 203 (2.6%) |
| Unknown/refused to say | 84 (1.8%) | 91 (2.7%) | 175 (2.2%) |
| Missing | 1 | 2 | 3 |
| **Fruit juices** | | | |
| None per week | 2,745 (60.2%) | 2,172 (65.6%) | 4,917 (62.5%) |
| One to two times a week | 1,458 (32.0%) | 942 (28.4%) | 2,400 (30.5%) |
| Three to four times a week | 225 (4.9%) | 105 (3.2%) | 330 (4.2%) |
| Five or more times a week | 118 (2.6%) | 78 (2.4%) | 196 (2.5%) |
| Refused to say | 13 (0.3%) | 15 (0.5%) | 28 (0.4%) |
| Missing | 1 | 2 | 3 |
| **Fizzy sugar sweetened beverages** | | | |
| None per week | 3,169 (69.5%) | 2,115 (63.9%) | 5,284 (67.1%) |
| One to two times a week | 1,227 (26.9%) | 1,020 (30.8%) | 2,247 (28.5%) |
| Three to four times a week | 122 (2.7%) | 131 (4.0%) | 253 (3.2%) |
| Five or more times a week | 23 (0.5%) | 32 (1.0%) | 55 (0.7%) |
| Refused to say | 18 (0.4%) | 14 (0.4%) | 32 (0.4%) |
| Missing | 1 | 2 | 3 |
| **Candy** | | | |
| None per week | 2,506 (55.0%) | 2,082 (62.9%) | 4,588 (58.3%) |
| One to two times a week | 1,624 (35.6%) | 971 (29.3%) | 2,595 (33.0%) |
| Three to four times a week | 299 (6.6%) | 174 (5.3%) | 473 (6.0%) |
| Five or more times a week | 115 (2.5%) | 70 (2.1%) | 185 (2.4%) |
| Refused to say | 15 (0.3%) | 15 (0.5%) | 30 (0.4%) |
| Missing | 1 | 2 | 3 |
| **Biscuits** | | | |
| None per week | 3,215 (70.5%) | 2,454 (74.1%) | 5,669 (72.0%) |
| One to two times a week | 1,135 (24.9%) | 707 (21.3%) | 1,842 (23.4%) |

| Characteristic | Female, n(%), 4560 (58%) | Male, n(%), 3314 (42%) | Total, n(%), 7874 (100%) |
|---|---|---|---|
| Three to four times a week | 140 (3.1%) | 107 (3.2%) | 247 (3.1%) |
| Five or more times a week | 55 (1.2%) | 28 (0.8%) | 83 (1.1%) |
| Refused to say | 14 (0.3%) | 16 (0.5%) | 30 (0.4%) |
| Missing | 1 | 2 | 3 |
| **Chocolate** | | | |
| None per week | 3,767 (82.6%) | 2,796 (84.4%) | 6,563 (83.4%) |
| One to two times a week | 671 (14.7%) | 427 (12.9%) | 1,098 (13.9%) |
| Three to four times a week | 87 (1.9%) | 57 (1.7%) | 144 (1.8%) |
| Five or more times a week | 20 (0.4%) | 17 (0.5%) | 37 (0.5%) |
| Refused to say | 14 (0.3%) | 15 (0.5%) | 29 (0.4%) |
| Missing | 1 | 2 | 3 |
| **Crisps** | | | |
| None per week | 2,072 (45.4%) | 1,760 (53.1%) | 3,832 (48.7%) |
| One to two times a week | 1,847 (40.5%) | 1,178 (35.6%) | 3,025 (38.4%) |
| Three to four times a week | 456 (10.0%) | 258 (7.8%) | 714 (9.1%) |
| Five or more times a week | 168 (3.7%) | 103 (3.1%) | 271 (3.4%) |
| Refused to say | 16 (0.4%) | 13 (0.4%) | 29 (0.4%) |
| Missing | 1 | 2 | 3 |
| **Fries/chips** | | | |
| None per week | 3,036 (66.6%) | 2,232 (67.4%) | 5,268 (66.9%) |
| One to two times a week | 1,254 (27.5%) | 884 (26.7%) | 2,138 (27.2%) |
| Three to four times a week | 204 (4.5%) | 134 (4.0%) | 338 (4.3%) |
| Five or more times a week | 50 (1.1%) | 47 (1.4%) | 97 (1.2%) |
| Refused to say | 15 (0.3%) | 15 (0.5%) | 30 (0.4%) |
| Missing | 1 | 2 | 3 |
| **Makoenya (traditional fried bread bun)** | | | |
| None per week | 2,260 (49.6%) | 1,638 (49.5%) | 3,898 (49.5%) |
| One to two times a week | 1,802 (39.5%) | 1,350 (40.8%) | 3,152 (40.0%) |
| Three to four times a week | 366 (8.0%) | 218 (6.6%) | 584 (7.4%) |
| Five or more times a week | 117 (2.6%) | 92 (2.8%) | 209 (2.7%) |
| Refused to say | 14 (0.3%) | 14 (0.4%) | 28 (0.4%) |
| Missing | 1 | 2 | 3 |
| **Meat** | | | |
| None per week | 1,952 (42.8%) | 1,303 (39.3%) | 3,255 (41.4%) |
| One to two times a week | 2,024 (44.4%) | 1,481 (44.7%) | 3,505 (44.5%) |
| Three to four times a week | 458 (10.0%) | 388 (11.7%) | 846 (10.7%) |
| Five or more times a week | 110 (2.4%) | 128 (3.9%) | 238 (3.0%) |
| Refused to say | 15 (0.3%) | 12 (0.4%) | 27 (0.3%) |
| Missing | 1 | 2 | 3 |
| **Smoking status** | | | |

| Characteristic | Female, n(%), 4560 (58%) | Male, n(%), 3314 (42%) | Total, n(%), 7874 (100%) |
|---|---|---|---|
| Current Smoker | 1,015 (22.3%) | 1,579 (47.9%) | 2,594 (33.1%) |
| Never Smoker | 3,500 (77.0%) | 1,657 (50.2%) | 5,157 (65.8%) |
| Ex-smoker quit smoking less than 5 years ago | 21 (0.5%) | 35 (1.1%) | 56 (0.7%) |
| Ex-smoker quit smoking 5 years ago or longer | 8 (0.2%) | 28 (0.8%) | 36 (0.5%) |
| Missing | 16 | 15 | 31 |
| **Frequency of alcohol consumption** | | | |
| Every day/ 7 days per week | 71 (1.6%) | 158 (4.8%) | 229 (2.9%) |
| 5 to 6 days per week | 43 (0.9%) | 102 (3.1%) | 145 (1.8%) |
| 3 to 4 days per week | 93 (2.0%) | 183 (5.5%) | 276 (3.5%) |
| 1 to 2 days per week | 287 (6.3%) | 505 (15.3%) | 792 (10.1%) |
| 1 to 3 days per month | 139 (3.1%) | 245 (7.4%) | 384 (4.9%) |
| Less than 1 to 3 days per month | 180 (4.0%) | 195 (5.9%) | 375 (4.8%) |
| Never | 3,729 (82.1%) | 1,914 (58.0%) | 5,643 (71.9%) |
| Missing | 18 | 12 | 30 |
| **First-degree relative with hypertension** | 976 (21.4%) | 374 (11.3%) | 1,350 (17.1%) |
| **First-degree relative with diabetes** | 257 (5.6%) | 90 (2.7%) | 347 (4.4%) |
| **History of myocardial infarction** | 62 (1.4%) | 11 (0.3%) | 73 (0.9%) |
| **History of chronic kidney disease** | 16 (0.4%) | 5 (0.2%) | 21 (0.3%) |
| **Current or previous TB diagnosis** | 169 (3.7%) | 238 (7.2%) | 407 (5.2%) |
| **Severe vision impairment/blindness** | 35 (0.8%) | 7 (0.2%) | 42 (0.5%) |
| **Peripheral arterial disease** | 27 (0.6%) | 5 (0.2%) | 32 (0.4%) |
| **Heart failure** | 32 (0.7%) | 5 (0.2%) | 37 (0.5%) |
| **History of stroke** | 20 (0.4%) | 13 (0.4%) | 33 (0.4%) |
| **History of neuropathy** | 39 (0.9%) | 8 (0.2%) | 47 (0.6%) |
| **Chronic lung disease (COPD/asthma)** | 16 (0.4%) | 11 (0.3%) | 27 (0.3%) |
| **Diabetic foot syndrome** | 6 (0.1%) | 1 (0.0%) | 7 (0.1%) |
| **Self-reported HIV status** | | | |
| HIV positive | 782 (17.1%) | 406 (12.3%) | 1,188 (15.1%) |
| Tested negative ≤ 12 months ago | 2,435 (53.4%) | 1,503 (45.4%) | 3,938 (50.0%) |
| Tested negative > 12 months ago | 846 (18.6%) | 684 (20.6%) | 1,530 (19.4%) |
| Unknown | 453 (9.9%) | 695 (21.0%) | 1,148 (14.6%) |
| Refused to say | 44 (1.0%) | 26 (0.8%) | 70 (0.9%) |
| **ART intake among those HIV positive (n=1188)** | | | |
| Yes | 771 (98.6%) | 398 (98.0%) | 1,169 (98.4%) |
| No | 11 (1.4%) | 8 (2.0%) | 19 (1.6%) |
| Missing | 3,778 | 2,908 | 6,686 |
| **Lipid lowering medication** | | | |
| Yes | 17 (0.4%) | 3 (0.1%) | 20 (0.3%) |
| No | 4,543 (99.6%) | 3,311 (99.9%) | 7,854 (99.7%) |
| **Aspirine** | | | |

| **Characteristic** | **Female, n(%)**, 4560 (58%) | **Male, n(%)**, 3314 (42%) | **Total, n(%)**, 7874 (100%) |
|---|---|---|---|
| Yes | 386 (8.5%) | 103 (3.1%) | 489 (6.2%) |
| No | 4,174 (91.5%) | 3,211 (96.9%) | 7,385 (93.8%) |

*Supplementary table 2 Overview of complete individual level demographic, clinical and behavioural risk factor data of adult participants.*
